# Statistical analysis plan (SAP): Risk of revision for different types of cup fixation in elderly patients with primary hip osteoarthritis. A study of XXX,XXX primary total hip arthroplasties from the Nordic Arthroplasty Register Association (NARA)

**DOI:** 10.1101/2024.05.26.24307935

**Authors:** Roshan Latifi, Simon Kornvig, Maziar Mohaddes Ardebili, Claus Varnum, Søren Overgaard

## Abstract

Uncemented fixation of THA components are increasingly used. Although cemented femoral component fixation in total hip arthroplasty has shown better results in older patients, less is known about the cup side.

## 2: Introduction

### 2.1 Describe briefly background, research questions and rationale behind the study

It is debated wether uncemented or cemented cup fixation should be used in older patients in total hip arthroplasty.

### 2.2 Describe briefly objectives and/or hypotheses

Overall aim:

The overall aim was to compare the risk of THA revision with cemented and uncemented acetabular cups.

The overall aim will be divided into 3 sub-aims:

1. Compare the risk of cup revision due to any reason after THA with cemented vs. uncemented acetabular cups
  1.1 Perform analysis 1) stratified by sex, 5-year age groups and type of bearing (PE vs. XLPE)
  1.2 Perform analysis 1) the first 2 years and the subsequent years after surgery
2. Compare the overall cause-specific risk of cup revision after THA with cemented vs. uncemented acetabular cups
  2.1 Perform analysis 2) stratified by sex, 5-year age groups and type of bearing (PE vs. XLPE)
  2.2 Perform analysis 2) the first 2 years and the subsequent years after surgery
3. Compare the risk of cup and/or stem revision to any reason after THA with cemented vs. uncemented acetabular cups
  3.1 Perform analysis 3) stratified by sex, 5-year age groups and type of bearing (PE vs. XLPE)
  3.2 Perform analysis 3) the first 2 years and the subsequent years after surgery

## Section 3: Study methods

### 3.1 Study design

Describe type of study (i.e. experimental/observational, parallel group/cross over, singlecenter/multicenter ect.) and describe briefly interventions

International register-based multicenter study

### 3.2 Randomization details (if applicable)

Describe randomization i.e. allocation ratio, potential factors randomization will be stratified for and describe how and when randomization will be performed

N.A.

### 3.3 Sample size

Describe calculation of sample size or reference to sample size calculation in study protocol

Approx. XXX,XXX THA procedures performed between 1995-2019

### 3.4 Hypotheses framework

Describe hypotheses framework i.e. superiority, equivalence or noninferiority hypothesis testing and which group comparisons will be analysed

N.A (two-sided testing)

### 3.5 Statistical interim analyses and stopping guidelines (if applicable)

Describe how and when interim analyses will be performed, and potential planned adjustment of significance level due to interim analyses. Describe guidelines for stopping the trial early.

N.A.

### 3.6 Timing of outcome assessments and follow-up

Describe time points at which outcomes/covariates will be measured (consider a figure to visualize the time windows of measurements – see appendix)

N.A. (register-based follow-up)

### 3.7 Timing of final analysis

i.e. all outcomes analysed collectively or analyses performed according to planned follow-ups

Collectively

## Section 4: Statistical principles and protocol deviations

### 4.1 Confidence intervals and P-values

Specification of level of statistical significance and confidence intervals to be reported. Describe, if relevant, rationale for adjustment for multipel testing and how type 1 error will be controlled for 95% CI will be applied

P-values <0.05 will be considered statistically significant. All hypothesis-testings will be two-sided.

In case of multiple testing, Holm-Bonferroni correction will be applied to control type 1 error rate.

### 4.2 Adherence/compliance and protocol deviations

Define adherence/compliance and how this is assessed in the study. Define protocol deviations and which protocol deviations will be summarized and presented

N.A.

### 4.3 Analysis populations

Define analysis population i.e. intention-to-treat, per-protocol, complete case, safety population

Complete-case

## Section 5: Study population

### 5.1 Screening (if applicable)

Describe screening data to determine eligibility (i.e. scoring and scales)

N.A.

### 5.1 Eligibility

Summarize in- and exclusion criteria

#### Inclusion

- Primary THA during January 1, 1995 to December 31, 2018.
- Performed on the indication primary osteoarthritis (coded 1), idiopatic femoral head necrosis/Perthes disiase (coded 5, 12, 13), developmental dysplasia of the hip (coded 10) and epifysiolysis capitis femoris (coded 11, 13)

#### Exclusion

- Age < 65 years
- Inverse hybrid
- Dual mobility cup
- Resurfacing (variable Fixtype)
- Implant used < 50 times in each country
- Missing data on confounders
- Patients registred with negative survival years
- Second THA in case of bilateral

### 5.2 Recruitment and flow chart

Specification of steps in the recruitment process i.e. enrollment, screening allocation for use in flow chart (see appendix)

**Roshan** will draft the flow-chart – see appendix for the STROBE template.

Simon will provide the numbers.

### 5.3 Withdrawal/loss to follow-up

Specification on how reason and timing of withdrawal or loss to follow-up will be recorded and presented (i.e. in the flow chart – see appendix)

Loss to follow-up may not have been registred in case of immigration etc., however survived years has been calculated as time from primary surgery date to death or end of follow up (December 31, 2020)

### 5.4 Baseline patient characteristics

List of baseline characteristics and how these data will be descriptively summarized in a “Table 1” (see appendix)

**Roshan** is responsible for drafting the tables – see appendix for suggestions.

Simon will provide the numbers.

## Section 6: Analysis

### 6.1 Exposure and outcome definitions

Describe details on exposure i.e. assessment, definitions, units and thresholds or the intervention/treatment under study.

List and describe details on primary and secondary outcomes i.e. definition of outcome and timing, specific clinical measurements and units (i.e. mmol/mol) or any calculation or transformation of data to derive the outcome (i.e. sum score, change from baseline, logarithm, quality-of-life scoring algorithm)

#### Exposure

All analyses will compare patients with cemented cups vs. uncemented acetabular cups. The exposure groups will be defined as:

Cemented = fixtype “cemented” (coded 1) Uncemented = fixtype “uncemented” (coded 2) and “hybrid” (3)

#### Outcome

Sub-aim 1) and 2) will consider cup revision defined as surgery procedure “Both cup and stem replaced”, “Only cup replaced” and “Definitive extraction (Girdlestone)”.

For sub-aim 2, cause-specific revision will be defined as Dislocation (coded as RevCause 5), Aseptic Loosening (RevCause 1) and Others (RevCause 2,3,7,9 and missing).

Sub-aim 3) will consider both cup and/or stem revision defined as the above-mentioned and in addition “Only stem replaced”, “others” and “missing”.

Only first revision will be considered.

### 6.2 Primary analysis methods

Describe in details which statistical methods will be used (i.e. regression), how treatment effects will be presented (i.e. which effect measure - OR, HR etc.) and if estimates will be adjusted for covariates (see appendix).

If analyses will be adjusted for covariates, describe how the sufficient adjustment set will be defined (i.e. using DAGs)

Describe methods used to check assumptions (i.e. normality, proportional hazards) behind the statistical models, and alternative methods if assumptions about distribution do not hold.

The following analysis will be performed according to the listed sub-aims:

#### 1 Compare the risk of cup revision due to any reason after THA with cemented vs. uncemented acetabular cups

Crude cumulative incidence function will be calculated and plotted using the Aalen-Johansen method for cemented cups and uncemented cups.

Risk of cup revision will be analysed using Cox regression models and reported as Hazard ratios (HR) with corresponding 95% CI. HRs will be reported crude and adjusted for potential confounders. A minimal adjustment set for the multivariate model is defined using Directed Acyclic Graphs (DAGs).

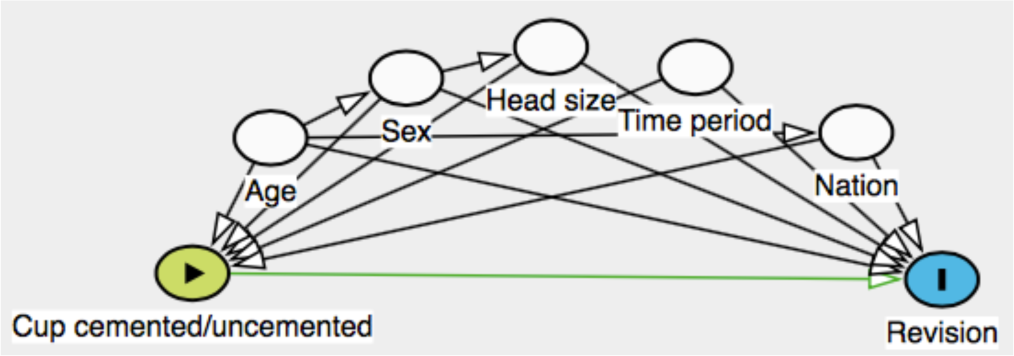

Minimal adjustment set: Age, sex, head size, time period and nation

Time period will be devided into 4 groups: 1995-1999, 2000-2005, 2006-2011, 2012-2018

Assumption of proportional hazards will be checked using log-log plots, and in case of uncertainty, supplied by Schoenfelds residuals.

1.1 Perform analysis 1) stratified by sex, 5-year age groups and type of bearing (PE vs. XLPE)
1.2 Perform analysis 1) the first 2 years and the subsequent years after surgery

Age groups will be defined with 5-year intervals accoding to age at primary surgery: 65-69 years, 70-74 years, 75-79 years and 80+

#### 2 Compare the cause-specific risk of cup revision after THA with cemented vs. uncemented acetabular cups

Cause-specific revision will be analysed in cause-specific Cox regression models taking into account competing risk from remaining revision causes and mortality. Estimates will be reported as HR with corresponding 95% CI. HRs will be reported crude and adjusted for potential confounders. A minimal adjustment set for the multivariate model is defined using DAGs (same as in 1)).

Assumptions will be checked according to 1).

2.1 Perform analysis 2) stratified by sex, 5-year age groups and type of bearing (PE vs. XLPE)
2.2 Perform analysis 2) the first 2 years and the subsequent years after surgery

Age groups will be defined with 5-year intervals accoding to age at primary surgery: 65-69 years, 70-74 years, 75-79 years and 80+

#### 3 Compare the risk of revision (cup and/or stem) due to any reason after THA with cemented vs. uncemented acetabular cups

Crude cumulative incidence function will be calculated and plotted using the Aalen-Johansen method for cemented cups and uncemented cups.

Risk of revision will be analysed using Cox regression models and reported as HR with corresponding 95% CI. HRs will be reported crude and adjusted for potential confounders.

A minimal adjustment set for the multivariate model is defined using DAGs (same as in 1)).

Assumptions will be checked according to 1).

3.1 Perform analysis 3) stratified by sex, 5-year age groups and type of bearing (PE vs. XLPE)
3.2 Perform analysis 3) the first 2 years and the subsequent years after surgery

Age groups will be defined with 5-year intervals accoding to age at primary surgery: 65-69 years, 70-74 years, 75-79 years and 80+

### 6.3 Additional analysis methods

Describe any planned sensitivity and subgroup analysis including how subgroups will be defined (see appendix).

#### Sensitivity analyses

Aim 1) and 3) will be analysed in Fine&Grey regression models taking into account the competing risk from mortality.

### 6.4 Missing data

Describe how missing data will be explored and which assumptions and methods will be used to handle missing data (i.e. multiple imputation)

Proportion and patterns of missing data will be explored, and in case of <5% missing data analyses will be performed as complete case. If the amount of missing data is comprehensive (>5%) or suggestive for biased estimates, appropriate methods of imputation will be applied depending on the patterns of missingness.

### 6.5 Harms (only applicable in experimental studies)

Describe the collection of safety data i.e. data on severity, expectedness, causality. Describe grouping and analyses planned i.e. incidence analyses on grade 3-4 events only.

N.A.

### 6.6 Statistical software

Specify statistical packages to be used for the analyses

STATA 17 or newer

## Data Availability

All data produced in the present study are available upon reasonable request to the authors

### Appendix: Figure and table templates

#### 5.2-3 Flow chart template for observational studies

**STROBE flow chart (4)**

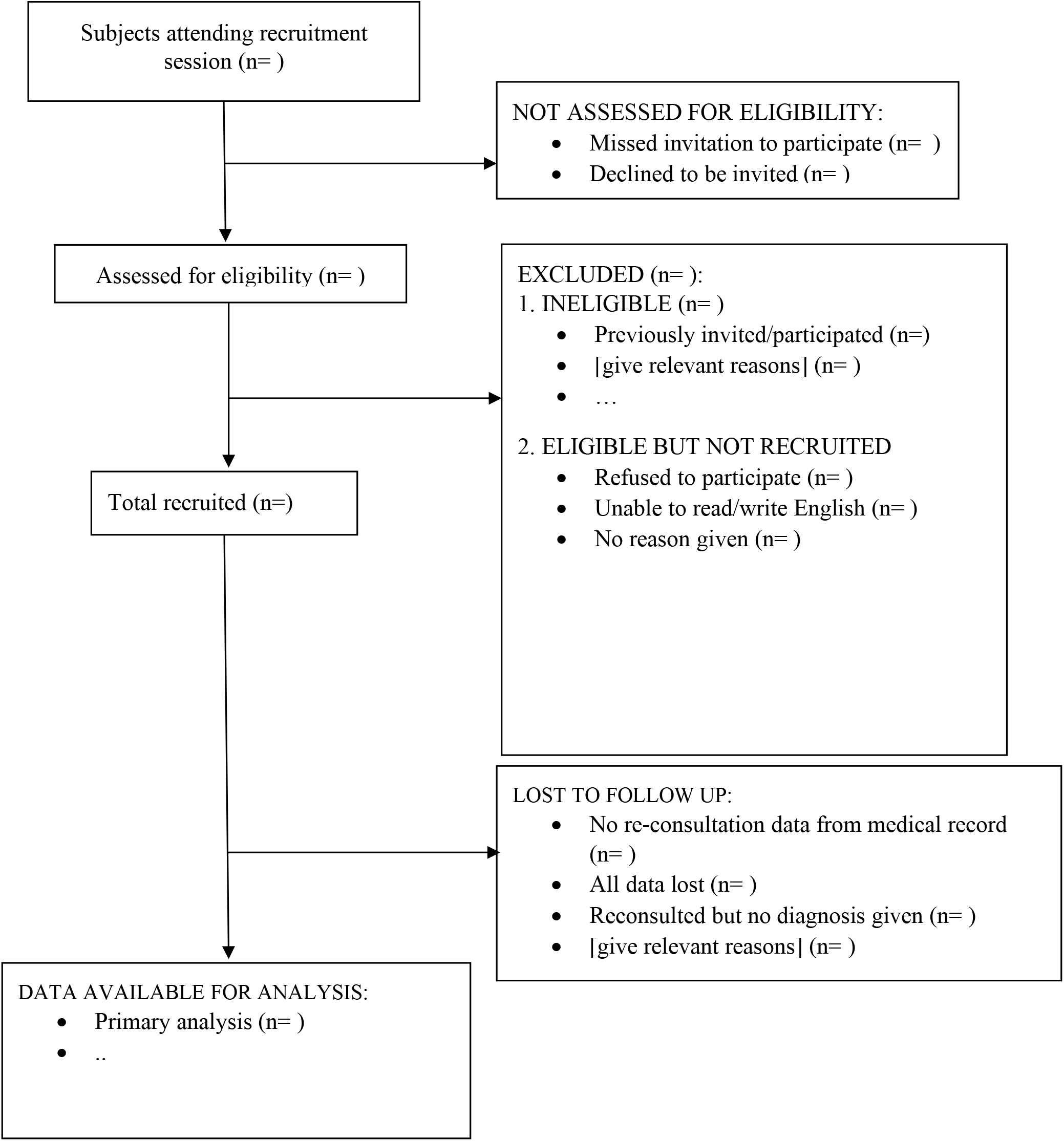

#### 5.4 Baseline table (“Table 1”) template

**Table 1:**
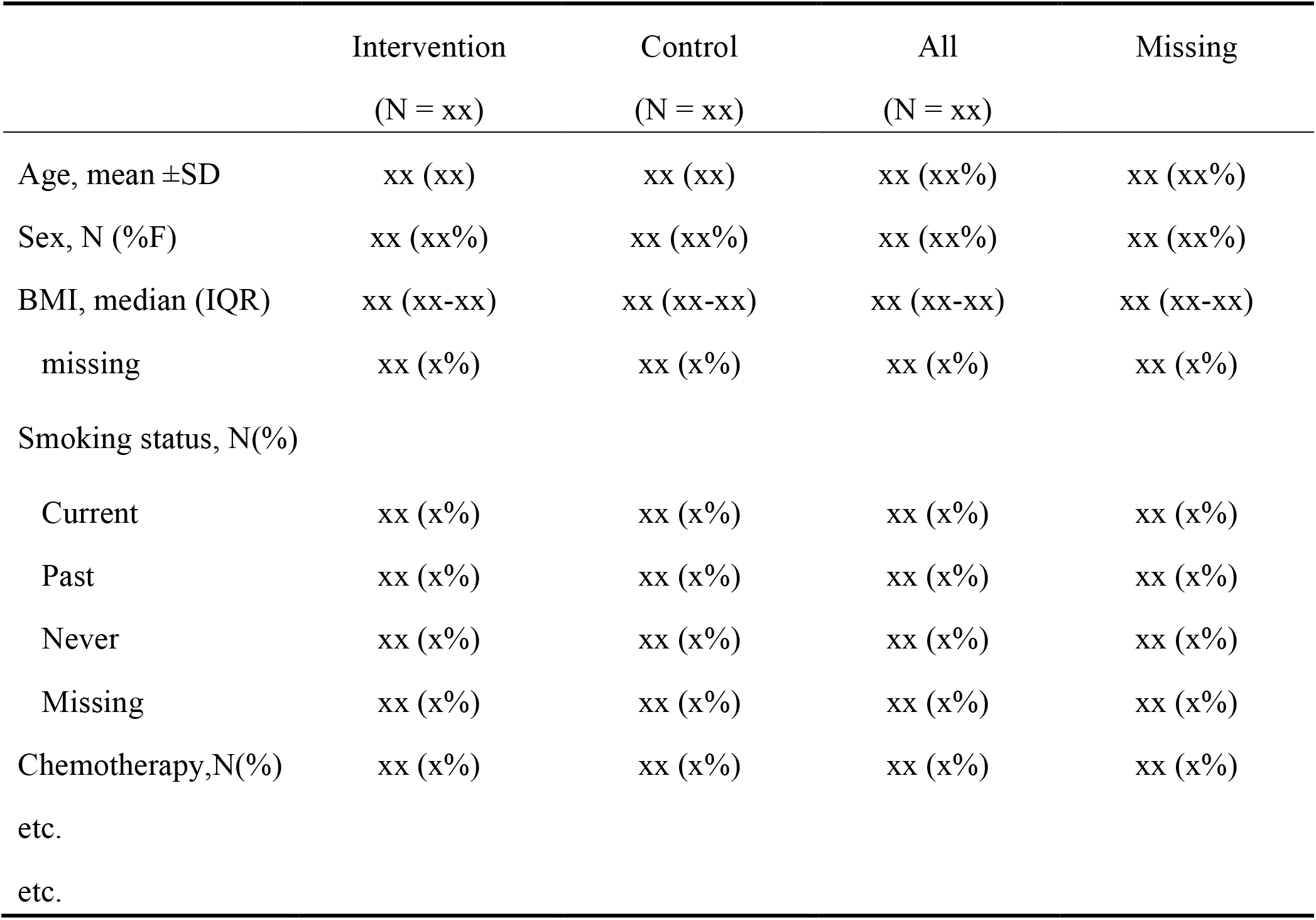
Characteristics of the study population.

|  | Intervention | Control | All | Missing |
| --- | --- | --- | --- | --- |
|  | (N = xx) | (N = xx) | (N = xx) |  |
| Age, mean ±SD | xx (xx) | xx (xx) | xx (xx%) | xx (xx%) |
| Sex, N (%F) | xx (xx%) | xx (xx%) | xx (xx%) | xx (xx%) |
| BMI, median (IQR) | xx (xx-xx) | xx (xx-xx) | xx (xx-xx) | xx (xx-xx) |
| missing | xx (x%) | xx (x%) | xx (x%) | xx (x%) |
| Smoking status, N(%) |  |  |  |  |
| Current | xx (x%) | xx (x%) | xx (x%) | xx (x%) |
| Past | xx (x%) | xx (x%) | xx (x%) | xx (x%) |
| Never | xx (x%) | xx (x%) | xx (x%) | xx (x%) |
| Missing | xx (x%) | xx (x%) | xx (x%) | xx (x%) |
| Chemotherapy,N(%) | xx (x%) | xx (x%) | xx (x%) | xx (x%) |
| etc. |  |  |  |  |
| etc. |  |  |  |  |

#### 6.2 Table example for reporting relative effect measures

**Table 2:** Relative effect measures on binary exposure and outcome.

|  | Crude RR (95% CI) | Adjusted RR (95% CI) | P-value |
| --- | --- | --- | --- |
| Primary exposure |  |  |  |
| No | 1 (ref) | 1 (ref) |  |
| Yes | xx (xx-xx) | xx (xx-xx) | xx |
| Secondary exposure |  |  |  |
| No | 1 (ref) | 1 (ref) |  |
| Yes | xx (xx-xx) | xx (xx-xx) | xx |

#### 6.3 Table example for reporting subgroup-analysis

**Table 3:**
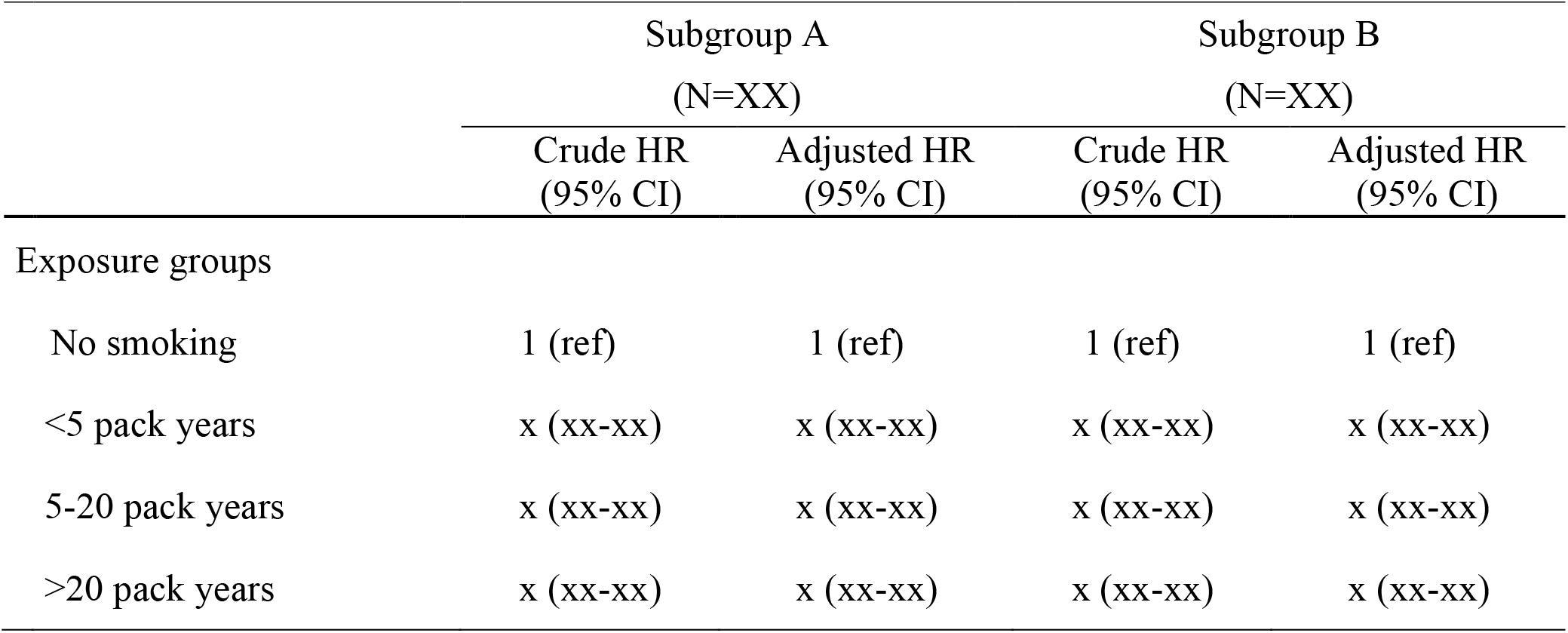
Subgroup-analysis on ordinal exposure and binary outcome reported with relative effect measure.

|  | Subgroup A<br>(N=XX) |  | Subgroup B<br>(N=XX) |  |
| --- | --- | --- | --- | --- |
|  | Crude HR<br>(95% CI) | Adjusted HR<br>(95% CI) | Crude HR<br>(95% CI) | Adjusted HR<br>(95% CI) |
| Exposure groups |  |  |  |  |
| No smoking | 1 (ref) | 1 (ref) | 1 (ref) | 1 (ref) |
| <5 pack years | x (xx-xx) | x (xx-xx) | x (xx-xx) | x (xx-xx) |
| 5-20 pack years | x (xx-xx) | x (xx-xx) | x (xx-xx) | x (xx-xx) |
| >20 pack years | x (xx-xx) | x (xx-xx) | x (xx-xx) | x (xx-xx) |

